# Spatial inequity in distribution of COVID-19 vaccination services in Aotearoa

**DOI:** 10.1101/2021.08.26.21262647

**Authors:** Jesse Whitehead, Polly Atatoa Carr, Nina Scott, Ross Lawrenson

## Abstract

**Aim:** This research examines the spatial equity, and associated health equity implications, of the geographic distribution of Covid-19 vaccination services in Aotearoa New Zealand.

**Method:** We mapped the distribution of Aotearoa’s population and used the enhanced-two-step-floating-catchment-method (E2SFCA) to estimate spatial access to vaccination services, taking into account service supply, population demand, and distance between populations and services. We used the Gini coefficient and both global and local measures of spatial autocorrelation to assess the spatial equity of vaccination services across Aotearoa. Additional statistics included an analysis of spatial accessibility for priority populations, including Māori (Indigenous people of Aotearoa), Pacific, over 65-year-olds, and people living in areas of high socioeconomic deprivation. We also examined vaccination service access according to rurality, and by District Health Board region.

**Results:** Spatially accessibility to vaccination services varies across Aotearoa, and appears to be better in major cities than rural regions. A Gini coefficient of 0.426 confirms that spatial accessibility scores are not shared equally across the vaccine-eligible population. Furthermore, priority populations including Māori, older people, and residents of areas with socioeconomic constraint have, on average, statistically significantly lower spatial access to vaccination services. This is also true for people living in rural areas. Spatial access to vaccination services, also varies significantly by District Health Board (DHB) region as does equality of access, and the proportion of DHB priority population groups living in areas with poor access to vaccination services. A strong and significant positive correlation was identified between average spatial accessibility and the Māori vaccination rate ratio of DHBs.

**Conclusion:** Covid-19 vaccination services in Aotearoa are not equitably distributed. Priority populations, with the most pressing need to receive Covid-19 vaccinations, have the worst access to vaccination services.

## Introduction

### Background

Aotearoa New Zealand’s (hereafter Aotearoa) elimination strategy in the fight against Covid-19 has put our country in a relatively good position internationally with few deaths, and low levels of active Covid-19 cases. ^1^ We are increasingly relying on a vaccination delivery programme to support an ongoing elimination strategy and to save morbidity and mortality. The recent outbreak of community transmission of the delta variant of Covid-19 highlights that, until the vaccination rollout is complete, there is a continued risk, particularly to priority populations. While vaccination helps reduce morbidity and mortality from infection, it does not stop transmission. Achieving the equitable and universal delivery of a vaccine is essential to help protect all residents of Aotearoa, and particularly communities that are at risk of severe Covid-19 outcomes. There are various ways of conceptualising ‘fairness’ and prioritising vaccine delivery, ^2^ ^3^ which in turn can have different impacts on Covid-19 related deaths, hospitalisations, and ICU admissions.^4^ Given the stark, persistent, and increasing health inequities in Aotearoa experienced particularly by Māori (the Indigenous people of Aotearoa),^5^ ^6^ it is essential to ensure that those at most risk of infection, and the members of the community most vulnerable to Covid-19 severity are prioritised for vaccine protection, and that every single person in the ‘team of 5 million’ has access to the vaccine. We have previously described priority populations as including Māori, Pacific, people aged 65 and over, people with comorbidities and those living in areas of high socioeconomic deprivation.^7^ A major priority should be to immunise people who might otherwise die if they contract Covid-19, while a secondary aim is to reduce admissions to hospital and to protect the health system.

It has been argued that governments should first allocate Covid-19 vaccines not only according to individuals’ risk of infection and underlying conditions, but that social vulnerabilities – such as socioeconomic status, high risk occupation, housing and living conditions, ethnicity, and other factors that limit access to healthcare – must also be considered. ^3^ It is particularly important to prioritise Indigenous populations.^8^ The transmission of Covid-19, and associated health impact is likely to be higher among Māori in Aotearoa New Zealand and Indigenous populations elsewhere. ^9^ Furthermore, in Aotearoa New Zealand it has been estimated that Covid-19 infection fatality rates could be 2.5 times higher for Māori than New Zealand Europeans, and 2 times higher for Pacific people.^10^ Health inequities between Māori and non-Māori are unjust, large, persistent, occur across the life course, and are well known by the Ministry of Health. Between 1992 and 2016 the Ministry of Health published 107 reports on Māori health and the disparity between Māori and non-Māori outcomes.^11^ Māori are disproportionately impacted by poorer access to the social determinants of health, including housing, quality healthcare, ^12^ and racism in the health system and wider society. ^13–17^

Achieving equitable immunisation necessarily involves balancing the logistical constraints of distributing and administering the time and temperature sensitive Covid-19 vaccine, with minimising barriers for those who wish to receive it. The vaccine roll-out in Aotearoa has taken a phased approach, grouping populations according to priority and risk. ^18^ Group 1 included with border and Managed Isolation and Quarantine (MIQ) workers (as well as their whānau and close contacts) who are consistently exposed to the greatest risk of infection. Group 2 targeted frontline health workers and people living in high-risk settings. Group 3 prioritised people aged 75 and older, people aged 65 and older, and those with underlying health conditions or disabilities. Group 4 includes the wider population of Aotearoa, and within Group 4 there has been a staged approach prioritising older age groups. The Pfizer vaccine has been offered across Aotearoa at a range of locations acting as vaccination services, including some general practitioner (GP) clinics and pharmacies. It has also been offered at a range of new ‘pop-up’ clinics at sports grounds, marae, and stadium mega-clinics. ^19^ The Ministry of Health has devolved decision making and the implementation of the vaccine rollout to District Health Boards (DHBs) who are responsible for the health of the populations in their region.

### Spatial accessibility and spatial equity

There is a risk that the white capture of resources and inequitable access to the vaccine that has been seen internationally,^3 20 21^ will occur in Aotearoa New Zealand. Access to health care in Aotearoa New Zealand is inequitable.^22 23–26^ This includes the inequitable geographic distribution of health services such as primary care services.^25^ ^27^ Spatial accessibility is not the only barrier to accessing healthcare, particularly for Māori,^28–31^ and the inequitable distribution of services is confounded by additional factors such as the (in)appropriateness, (un)availability, (in)acceptability and poorer quality care provided by many services. Māori and Pacific people report experiencing racism from healthcare providers,^13–17^ and are also disproportionately affected by cost and transport as barriers to accessing GP services.^32^ Barriers to accessing the vaccine disproportionately affecting those who are at the most risk of Covid-19 severity, such as people with underlying conditions and those aged over 65 years old, as well as Māori, Pacific and socioeconomically deprived communities which experience higher levels of chronic disease^22^ ^23^ will exacerbate key inequities. The Ministry of Health and DHBs should be aiming for an equitable and universal vaccine rollout. This must include ensuring a spatially equitable vaccine rollout which gives priority populations appropriately higher access to vaccination services and opportunities for vaccination. This paper examines the spatial equity of the vaccine rollout in Aotearoa with a particular focus on priority populations.

## Methodology

### Key steps

Four key steps were taken in the assessment of the spatial equity of Covid-19 vaccination services (see Figure 1).

**Figure 1:**
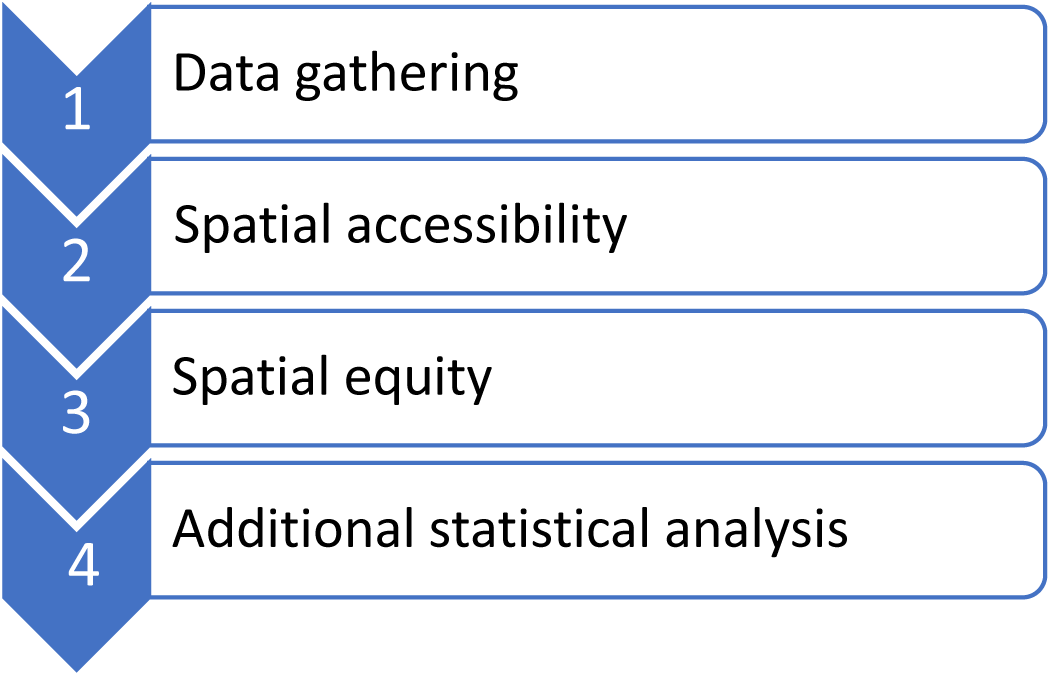
Key steps in assessing the spatial equity of Covid-19 vaccination services

### Data gathering

The population data used in this analysis is based on 2018 census data at the Statistical Area 1 (SA1) level. This includes information on the usually resident population, as well as the age and ethnicity of residents in each SA1.^33^ Ethnicity data in the 2018 census are not prioritised, so individuals who report multiple ethnicities are counted more than once.^34^ The population aged 15 and over was used to represent the ‘vaccine eligible’ population. At the time of analysis, the Pfizer population had only been approved in Aotearoa for people aged 16 and over. At the time of writing this has been updated to include children aged 12-16. Since the SA1 census dataset groups ages into 5-year-bands (e.g. 0-4, 5-9, 10-14, 15-19, etc) it was decided that the population aged 15 and over would be used for this analysis. It is important to note that this may be an underestimate of the total vaccine eligible population. The 2018 New Zealand Index of Socioeconomic Deprivation (NZDep18) information was accessed from the University of Otago.^35^ Geographic data include SA1 address-weighted centroids developed using the SA1 boundaries^36^ and address points^37^ datasets. The Geographic Classification for Health (GCH),^38^ a novel rurality classification specifically designed and developed for health research purposes was used to define rural and urban areas of Aotearoa. The road network layer used in this analysis was developed by Beere.^39^ Information of the location of COVID-19 vaccination services across Aotearoa was accessed on 18^th^ August 2021 from the Health Point^19^ website, which is a private company that provides health information – including information on medical services, and Covid-19 testing and vaccination services – on behalf of the health system. Vaccination services were linked to the Ministry of Health Facilities dataset^40^ which includes XY coordinates for all health facilities in Aotearoa. If vaccination services could not be located in the facilities dataset, their location was manually geocoded.

### Spatial accessibility

Geographic Information Systems (GIS) were used to quantitatively assess the spatial equity of COVID-19 vaccination services. The three steps to spatial equity analysis involve defining, estimating and quantifying spatial equity.^41^ Although spatial equity has a range of definitions that vary with context,^42^ it has been referred to as a fair distribution of resources relative to need.^43^ This recognises that in order to achieve equitable health outcomes, some populations with higher needs or underlying vulnerabilities should be prioritised and have better access to services.^6^ Similarly, there are a range of measures and techniques used to estimate the spatial accessibility of health services.^44^ The “Floating Catchment Area” (FCA) group of techniques estimate accessibility by considering service availability relative to population size and the distance between populations and services. FCAs calculate the ratio between the number of services and the size of populations within a defined catchment area and produce an accessibility score for each small area unit within a study area.^45^ The main advancement of the Enhanced-2-step-floating-catchment-area method (E2SFCA) is that it incorporates a distance decay function, which recognises that spatial access to services decreases for populations living further from the centre of a GP catchment. The E2SFCA is now considered the default spatial accessibility measure.^46^ This paper applied the E2SFCA method in ArcGIS^47^ to estimate accessibility to COVID-19 vaccination services in Aotearoa, using the 30 minute drivetime catchments originally proposed. ^48^ ^49^ The ArcGIS OD-Matrix was used to identify all SA1 centroids (and their associated resident populations) and vaccination services that were within 30 minutes’ drive from each other. In step 1, the OD-Matrix was used to calculate a supply-to-demand ratio for each vaccination service based on the total vaccine eligible population able to access it. In step 2, the ratio scores of vaccination services within reach of each SA1 centroid were then summed to give an accessibility score for each SA1 in Aotearoa. The Butterworth continuous distance-decay function (^50^ was applied at both steps.

### Spatial equity

Once overall levels of accessibility have been estimated, the Gini coefficient can be used to quantify equality. The Gini coefficient assesses the distribution of resources (such as income, or in this case, accessibility) across a population, and provides an equality score between 0 and 1, with 0 representing a perfectly equal distribution and 1 indicating a completely unequal distribution.^51^ To examine the equality of spatial access to COVID-19 vaccination services, the population weighted Gini coefficient was calculated in R ^52^ using the ACID package.^53^

Although the Gini coefficient gives an indication of whether the distribution of spatial accessibility to vaccination services is equal, it does not indicate whether such a distribution is *equitable*. For instance, in a system where the entire vaccine eligible population has the same level of access to vaccination services, access would be inequitable for priority populations. Therefore, it is important to examine which locations and populations have high or low levels of access to services. To identify whether there was any statistically significant clustering of spatial accessibility scores the Global Moran’s | measure of spatial autocorrelation was calculated in Arc GIS. Global Moran’s */* quantifies the degree of spatial clustering or dispersion of a value, in this case spatial accessibility, and determines whether it is statistically significant.^54^ Anselin’s Local Indicator of Spatial Autocorrelation Moran’s */* (LISA) was also calculated to map the locations of statistically significant clusters of high and low access.

### Additional statistical analysis

Additional statistical tests were undertaken to determine whether spatial access to vaccination services varied for priority populations – particularly for Māori, Pacific, older people, and those living in areas of high socioeconomic deprivation. Differences in spatial access to vaccination services between rural and urban areas of Aotearoa were also examined. To establish whether average spatial accessibility scores, as estimated by the E2SFCA, vary significantly for different population groups, independent-samples t-tests with the Bonferroni adjustment were calculated in R using the “ez” package.^55^ Statistical significance was defined as p<0.01. A one-way ANOVA and adjusted independent-samples t-tests were also used to determine whether there was a statistically significant difference in the average spatial accessibility scores for each DHB region. The proportion of each priority population group living in areas with poor access to vaccination services (Quintile 5) was also calculated for each DHB region.

## Results

### Spatial accessibility

Through the Health Point website we identified 447 vaccination services across Aotearoa, of which 212 (47%) were identified as GP clinics, 91 (20%) were pharmacies, 50 (11%) appeared to be DHB-run dedicated vaccination centres, and 28 (6%) appeared to be iwi led or run by Māori or Pacific providers. Figure 2 shows the locations of these vaccination services and indicates the geographic distribution of spatial accessibility scores across Aotearoa as estimated by the E2SFCA method. Scores were sorted into quintiles, with Quintile 1 (Q1 – best access) represented in light red and Quintile 5 (Q5 – worst access) in dark red. Figure 2 suggests that while access to COVID-19 vaccination services in large cities is generally good, there are large parts of rural Aotearoa with poor access. Of the major centres, Ōtautahi appears to have the worst access, while Te Whanganui-a-Tara and Ōtepoti have good levels of access to vaccination clinics.

**Figure 2:**
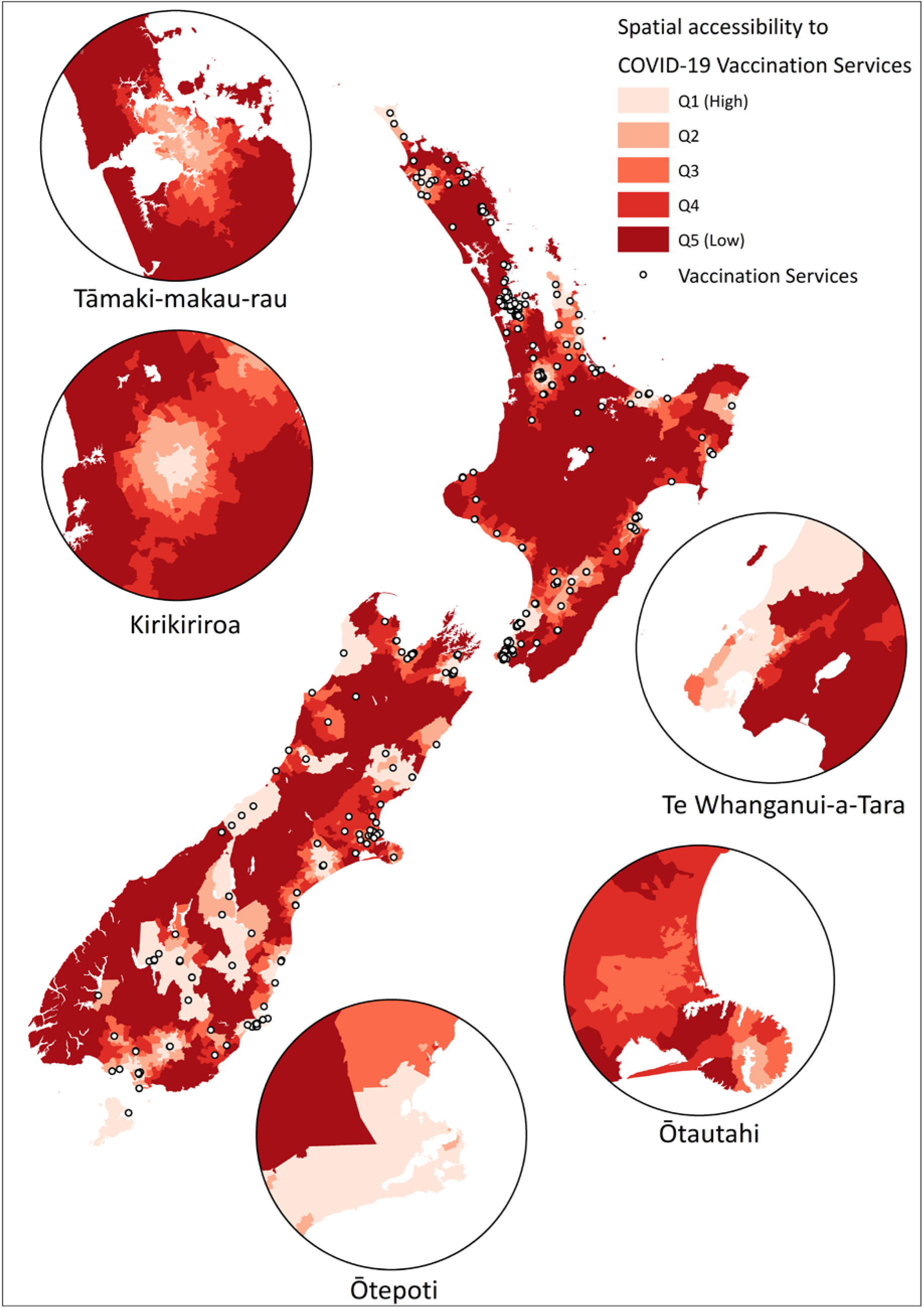
Spatial accessibility of Covid-19 vaccination services in Aotearoa

### Spatial equity

#### Gini coefficient

The Gini coefficient for the distribution of spatial access to vaccination services across Aotearoa was 0.426, suggesting an unequal distribution of vaccination services.

#### Spatial autocorrelation

Global Moran’s */* returned a statistically significant result (*I =* 0.349, p<0.00), indicating that spatial accessibility scores are clustered. The LISA analysis results are shown in Figure 3, and indicates where those clusters are. Dark green represents high-high clusters, which are statistically significant clusters of areas of high accessibility surrounded by other high access areas. Areas in light green are high-low outliers, which have high accessibility but are surrounded by areas with low access. Similarly, the dark blue regions represent low-low clusters, while light blue areas are low-high outliers. Figure 3 highlights that clusters of high accessibility tend to be in major cities, while rural and remote areas of Aotearoa have clusters of poor access to vaccination services.

**Figure 3:**
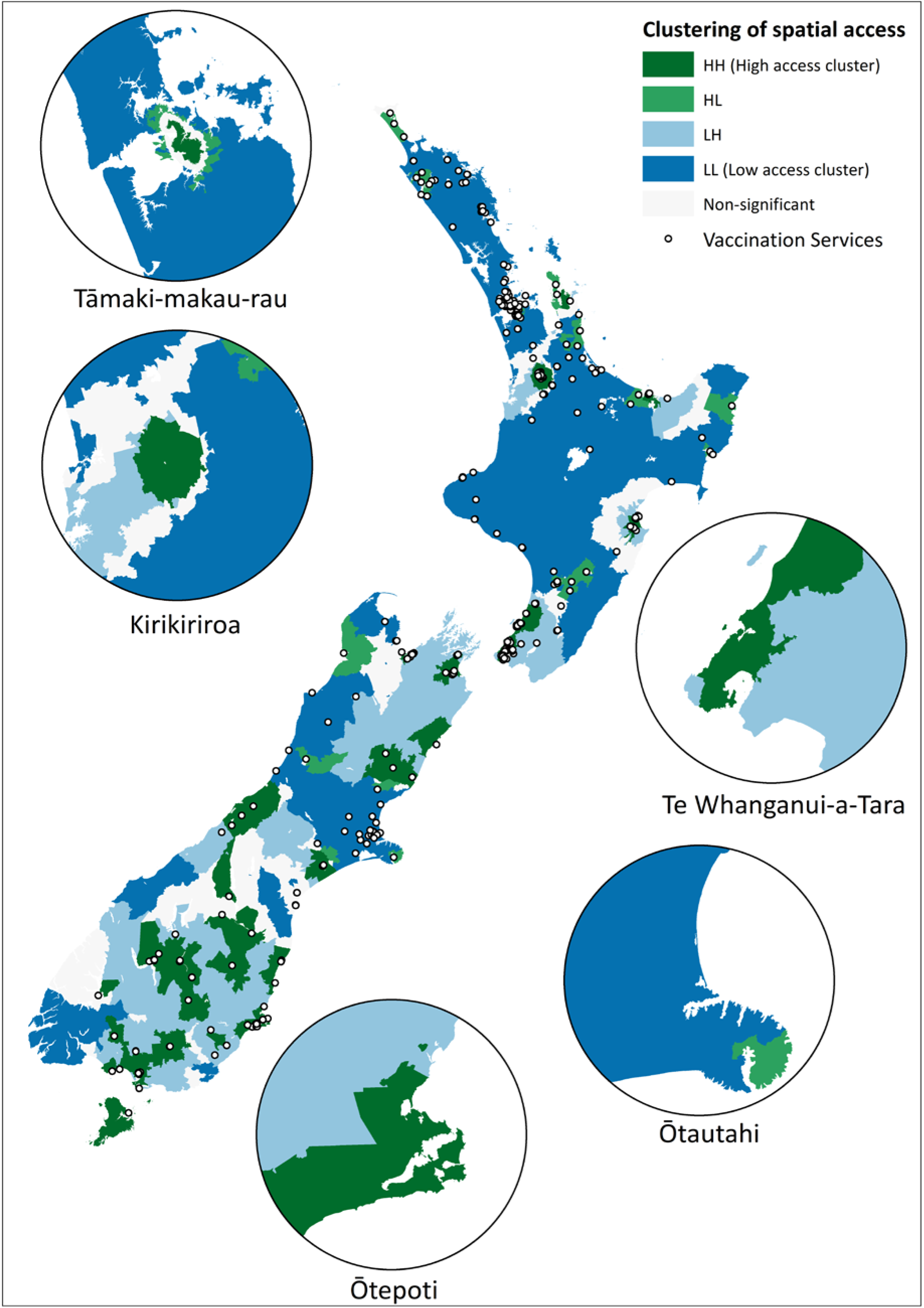
Spatial clustering of Covid-19 vaccination services in Aotearoa

### Additional statistical analysis

Further statistical analysis of spatial accessibility scores indicates that differences in access to vaccination services negatively affect priority populations, and are therefore inequitable. Table 1 displays the average spatial accessibility scores of different types of neighbourhoods. Spatial accessibility scores reflect the level of spatial access that people living in a neighbourhood have to vaccination services, with higher scores indicating better spatial access to vaccination services as estimated by the E2SFCA which calculates scores based on levels of supply, population demand, and distance to services. Spatial accessibility scores across Aotearoa ranged from 0.0 to 382.4, with an average score of 13.6, a median score of 11.1, and an interquartile range of 12.2.

**Table 1:** Differences in average spatial accessibility by neighbourhood type

| Neighbourhood type | Average spatial accessibility |
| --- | --- |
| High % Māori | 11.7 |
| Low % Māori | 14.8 |
| High % Pacific | 14.4 |
| Low % Pacific | 13.4 |
| High % over 65 years | 12.9 |
| Low % over 65 years | 14.1 |
| NZDep Q1 | 14.4 |
| NZDep Q2 | 13.8 |
| NZDep Q3 | 13.2 |
| NZDep Q4 | 13.3 |
| NZDep Q5 | 13.2 |
| Rural | 11.8 |
| Urban | 14.1 |
| Total Aotearoa | 13.6 |

Independent-samples t-tests revealed statistically significant differences in the average spatial accessibility scores of some types of neighbourhoods. Areas with a higher proportion of Māori residents (>15%) had statistically significantly worse access to vaccination services than neighbourhoods with a lower proportion of Māori residents (*p*<.001). Similarly, neighbourhoods with a higher proportion of over 65-year-olds (>15%) had worse access to vaccine services than areas with a lower proportion of over 65-year-olds (*p*<.001). Conversely, neighbourhoods with a high proportion of Pacific residents (>8%) had, on average, slightly better access to vaccine services than areas with a low proportion of Pacific residents (*p*<.001). There was a statistically significant difference between average accessibility scores in areas of low compared to high socioeconomic deprivation. Wealthier neighbourhoods had better access to vaccination services than neighbourhoods in NZDep2018 quintiles 3-5 (*p*<.001). Rural residents also had, on average, worse access to vaccination services than residents of urban areas (*p<*.001).

There was also a statistically significant difference between average levels of spatial accessibility between DHBs as determined by a one-way ANOVA (*F*(20, 29667) = 995.6, *p*<.001). Table 2 shows, for each DHB region, the average spatial accessibility score, Gini coefficient, and the proportion of each priority population group (as well as the total eligible population) that is living in an area with poor spatial access (Q5) to vaccination services. Table 2 also displays vaccination Rate Ratios for Māori and Pacific residents of each DHB region, indicating the relative proportion of Māori and Pacific who have received 2 vaccine doses, as compared to non-Māori and non-Pacific residents. DHBs with higher levels of average spatial accessibility (e.g. Capital & Coast and Southern DHB) appear to have more equitable levels of vaccine uptake for Māori and Pacific people living in the region. A strong and statistically significant correlation was identified between DHBs’ average levels of spatial accessibility and vaccination rate ratios for Māori (*r*=.84, *p*<.001) but not Pacific (*r=*.39, *p*=.079). Weak and non-significant correlations were identified between DHBs’ Gini coefficients and vaccination rate ratios for Māori (*r*=−.17, *p*=.46) and Pacific (*r=*-.21, *p=*.36)

**Table 2:** Average spatial access, Gini coefficient and % of priority populations living in areas with poor access to vaccination services for each DHB region

| DHB region | Average Spatial Access | Gini | % Māori in low access areas | % Pacific in low access areas | % over 65 in low access areas | % Eligible in low access areas | Māori vaccine Rate Ratio‡ | Pacific vaccine Rate Ratio |
| --- | --- | --- | --- | --- | --- | --- | --- | --- |
| Auckland | 14.7 | 0.131 | 3.2 | 0.6 | 4.0 | 2.2 | 0.63^ | 0.77^ |
| Bay of Plenty | 7.6 | 0.392 | 25.3 | 25.2 | 23.1 | 26.3 | 0.52 | 1.04 |
| Canterbury | 11.1 | 0.247 | 3.3 | 1.2 | 3.3 | 3.4 | 0.78 | 0.96 |
| Capital and Coast | 31.6 | 0.089 | 0.0 | 0.0 | 0.0 | 0.0 | 1.04^^ | 1.29^^ |
| Counties Manukau | 13.4 | 0.208 | 8.8 | 1.4 | 10.9 | 8.3 | 0.63^ | 0.77^ |
| Hawke's Bay | 13.8 | 0.208 | 5.8 | 1.7 | 3.7 | 4.9 | 0.58 | 1.29 |
| Hutt Valley | 19.6 | 0.290 | 1.2 | 0.6 | 3.6 | 3.3 | 1.04^^ | 1.29^^ |
| Lakes | 2.1 | 0.237 | 99.3 | 99.5 | 99.4 | 99.5 | 0.48 | 0.69 |
| MidCentral | 13.3 | 0.191 | 3.0 | 1.3 | 3.9 | 4.1 | 0.61 | 0.65 |
| Nelson Marlborough | 15.9 | 0.233 | 4.7 | 2.1 | 5.0 | 5.1 | 0.61 | 1.47 |
| Northland | 2.6 | 0.478 | 88.6 | 90.8 | 93.0 | 92.6 | 0.51 | 0.56 |
| South Canterbury | 11.0 | 0.284 | 12.0 | 11.9 | 17.1 | 17.1 | 0.71 | 0.45 |
| Southern | 31.0 | 0.287 | 3.4 | 1.4 | 3.2 | 3.8 | 1.21 | 0.81 |
| Tairāwhiti | 11.3 | 0.157 | 11.2 | 4.8 | 6.9 | 8.7 | 0.55 | 0.69 |
| Taranaki | 5.6 | 0.136 | 10.8 | 5.7 | 7.3 | 10.1 | 0.60 | 0.99 |
| Waikato | 13.4 | 0.298 | 21.4 | 22.7 | 18.4 | 17.0 | 0.67 | 0.93 |
| Wairarapa | 4.6 | 0.197 | 39.5 | 40.9 | 31.6 | 35.1 | 0.56 | 1.09 |
| Waitemata | 4.1 | 0.358 | 53.0 | 41.0 | 59.4 | 56.8 | 0.63^ | 0.77^ |
| West Coast | 15.9 | 0.675 | 11.2 | 10.1 | 10.2 | 11.7 | 0.59 | 0.86 |
| Whanganui | 7.0 | 0.276 | 32.4 | 37.2 | 26.2 | 29.5 | 0.50 | 0.52 |
| Total Aotearoa | 13.6 | 0.426 | 25.3 | 11.4 | 20.0 | 19.6 | 0.64 | 0.86 |

‡Rate Ratios are reported by the Ministry of Health.<sup>56</sup> The data represented in this table was reported on 20<sup>th</sup> August 2021, and represents vaccination rates for Māori and Pacific, relative to rates for Non-Māori-non-Pacific as at 19<sup>th</sup> August 2021.
^ The Ministry of Health reports Rate Ratios for Auckland, Waitemata, and Counties Manukau DHBs together as "Auckland Metro"
^^ The Ministry of Health also reports Rate Ratios for Capital & Coast and Hutt Valley DHBs together as "Capital & Coast and Hutt Valley"

## Discussion

There are some limitations to this work. Although information on vaccination services listed on the Health Point website is likely to be accurate and reflective of the actual services available on 18^th^ August 2021, it will not reflect all of the additional clinics that have been set up since that date in response to the Covid-19 delta variant outbreak which began on 17^th^ August 2021. Our analysis does not include any vaccination services that are not listed on the Health Point website. While this limitation is beyond the control and scope of this paper, it does highlight the importance of strong public health intelligence, including the collection and maintenance of information on service delivery. Furthermore, it is important to note that our analysis has not included any information on the capacity of vaccination services, the availability of appointments, or the different service models that may be used by vaccination services. Likewise, it does not assess the wider domains of accessibility (such as the acceptability or services) beyond spatial access.

Despite these limitations, this analysis indicates that, as has been predicted previously,^7^ spatial access to vaccination services across Aotearoa is inequitable. It shows that, in particular, Māori, over 65-year olds, people living in areas of high socioeconomic constraint, and rural residents have worse access to vaccination services. Given the higher burden of disease, and likelihood of more severe outcomes of Covid-19 infection in these groups, it is essential to ensure that priority populations are able to be vaccinated as soon as possible. The Ministry of Health and DHBs have had the opportunity to work with priority communities - including Iwi Māori, Pacific, and rural communities - to ensure an equitable vaccination rollout. The location of vaccination services could have been proactively planned to target priority populations and maximise access opportunities for these groups. Our finding that more than two-thirds of vaccination services are run from health facilities such as GP clinics, pharmacies, and hospitals suggest that authorities have relied on current health services, regardless of their inequitable distribution already highlighted in the research literature.^7 25–27 31^ This decision appears to have resulted in a disproportionate burden to accessing a Covid-19 vaccination for older people, Māori, rural people, and residents of neighbourhoods with socioeconomic constraint, all groups who are at risk of severe outcomes from Covid-19 infection.

Furthermore, our finding of significantly lower spatial access to vaccination services for communities with a higher proportion of Māori residents, and that more than a quarter of Māori live in areas with low access to vaccination services, indicates structural racism in Aotearoa’s Covid-19 vaccination rollout. Current available data from the Ministry of Health^57^ shows that compared to non-Māori and non-Pacific overall vaccination rates are lower for Māori (Dose 1 = 0.67, Dose 2 = 0.69) and Pacific (Dose 1 = 0.91, Dose 2 = 0.92). This is particularly stark for younger Māori, where the rate ratios are 0.54 and 0.48 for Dose 1 and Dose 2 respectively. When considered alongside our results, this underlines that the Ministry of Health led Covid-19 vaccination rollout has failed Māori, and reinforces the urgent need for an independent Māori Health Authority, with a service commissioning mandate, to design and deliver effective and equitable services for Māori.

Our results also highlight significant variation in levels of access - and inequity in access - to vaccination services between DHBs. This is not surprising in and of itself, as localised decision making around the delivery of Covid-19 vaccinations has been devolved from the Ministry of Health to DHBs, which are likely to have followed different vaccine rollout plans, had different levels of partnership with Iwi, and relied on the existing distribution of health facilities (which already provide differing levels of access to services across DHBs). These differences in access and spatial equity between DHBs appears to be associated with the equity of vaccine uptake for Māori and Pacific. A strong and statistically significant correlation between average spatial accessibility and Māori vaccination rate ratios was identified. Capital & Coast DHB has both the highest average level and most even distribution of spatial access to vaccination services, and high relative vaccination rates for Māori and Pacific living in the region. Similarly, Southern DHB has high levels of average access to services, and the highest relative vaccination rate for Māori. On the other hand, DHBs which are providing low levels of spatial access to vaccination services, such as Lakes, Northland, Bay of Plenty and Whanganui all have low vaccination Rate Ratios for Māori (0.48 – 0.52).

This suggests that improving the spatial equity of Covid-19 vaccination services, by offering additional services in areas with high priority populations and low access to current vaccination services, will be important for improving the equity of vaccination uptake and protecting priority populations. It is important to note that the Ministry of Health has not yet reported vaccination rates according to area-level socioeconomic deprivation, or rural-urban status, and therefore the equity of the overall vaccination rollout for these groups is unknown. Furthermore, DHB-level vaccination rates are not readily available for these groups, and therefore the impact of differing levels of spatial access to vaccination services for these groups on vaccination uptake is also currently unknown. Before Aotearoa can consider loosening international border restrictions, or moving from an elimination to a suppression strategy, it will be essential to achieve high vaccination rates among the priority populations who would experience the most severe health outcomes from Covid-19 infection. Our findings emphasise the importance of and need for national strategies that make use of both geospatial and public health intelligence to guide a national vaccination rollout – and in a wider sense, the equitable delivery of health services in general.

## Data Availability

The spatial accessibility data referred to in the manuscript is available on request. Other data used was open source and is freely available from the sources listed.

